# Domestic Violence across 30 Countries in Africa

**DOI:** 10.1101/2020.05.19.20107029

**Authors:** Anna E. Ssentongo, Emily S. Heilbrunn, Paddy Ssentongo, Dan Lin, Yanxu Yang, Justin Onen, Joshua P. Hazelton, John S. Oh, Vernon M. Chinchilli

**Affiliations:** Department of Public Health Sciences, Penn State College of Medicine and Milton S. Hershey Medical Center, Hershey, Pennsylvania, United States of America; Department of Surgery, Division of Trauma Surgery, Penn State College of Medicine and Milton S. Hershey Medical Center, Hershey, Pennsylvania, United States of America; Center for Neural Engineering, Department of Engineering, Science and Mechanics, The Pennsylvania State University, Pennsylvania, United States of America; CURE Children’s Hospital of Uganda, Mbale, Uganda

## Abstract

**Introduction:** Domestic violence is a prevalent global health issue that causes incredibly adverse consequences for an individual’s physical and psychological health. The rates of physical and sexual violence in developing countries are thought to be some of the highest in the world, where up to 44% of women in Sub-Saharan Africa have experienced domestic violence. However, this has not been explored systematically. We present the first study to estimate the incidence and prevalence of physical and sexual violence across Africa.

**Methods:** Poisson meta-regression analysis on demographic health survey data from 482,670 women from 442,507 households in 30 countries across Africa. Hotspot analysis using the Getis-ORD Gi approach at the sub-regional level.

**Results:** Of 482,670 women, those who were divorced or separated were approximately 7 times more likely to experience physical violence and 6 times more likely to experience sexual violence compared to those who were never married (Risk Ratio: RRs:7.35, 95%CI 7.16-7.54 and 5.89, 95%CI 5.75-6.02 respectively). Likewise, wealth index and education level were inversely related to the incidence and prevalence of sexual and physical violence. Hotspots of sexual and physical violence were identified in Congo and surrounding areas.

**Conclusions:** Interventions should be designed to address the high levels of physical and sexual violence in Congo and surrounding areas, especially in those who are less educated, have lower wealth indices, and are divorced or separated.

**Key Questions:** *What is already known?:* Experiencing domestic violence can increase the likelihood of experiencing poorer health outcomes. Previous research has shown that the rates of domestic violence is higher in developing countries, specifically in countries located in sub-Saharan Africa.

*What are the new findings?:* This is the first study to estimate the incidence and prevalence of physical and sexual violence across regions in Africa. Geospatial analysis illustrated hot-spots near Congo that have a higher prevalence of domestic violence. This study also solidified the association between income, education, and marital status, and domestic violence. Those who are less educated and are of lower socioeconomic status are more likely to experience domestic violence in comparison to their educated, wealthy counterparts.

*What do the new findings imply?:* Our findings further promote the importance of tailoring appropriate interventions that strive to reduce domestic violence across Africa, while also protecting domestic violence victims against the adverse psychological and physical health outcomes that may occur as a result of their exposure.

## Introduction

Domestic violence (DV), as defined by the World Health Organization, is any form of a behavior that causes psychological, physical, or sexual harm to an intimate partner.[1] Experiencing domestic violence of any form has the potential to cause incredibly adverse consequences for an individual’s physical and psychological health. Prior research supports the notion that victims of domestic violence may experience poorer physical and mental health outcomes in comparison to those who have not experienced domestic violence.[2] The psychological consequences of experiencing domestic violence have been explored greatly in research. Depression, anxiety, post-traumatic stress disorder (PTSD), suicidal ideation, and self-injury are just a few of the many psychological consequences of domestic violence.[2] Domestic violence also has major implications for both acute and chronic health conditions, with victims having a 60% higher rate of total health problems in comparison to individuals who have never experienced any form of abuse (incidence rate ratio: 1.58; 95% CI 1.34-1.86).[3] Research has found that those who have experienced domestic violence are more likely to report chronic conditions, including cardiovascular disease, high blood pressure, elevated cholesterol, and more.[4] Experiencing domestic violence also increases a victim’s likelihood of participating in risky health behaviors, such as smoking and binge drinking.[4]

Domestic violence is an extremely prevalent global health problem, with approximately one in three women experiencing some form of DV during their lifetime in the United States.[5] Approximately 10 million individuals within the United States experience some form of physical abuse annually.[6] The average global prevalence of domestic violence is approximately 30%.[7] European countries (prevalence: 25.4%, 95% CI 20.9-30.0) and Western Pacific regions (prevalence: 24.6%; 95% CI 20.1-29.0) report rates of domestic violence that are significantly lower than the global average.[6] The growing prevalence of domestic violence across the globe is extremely concerning, for the economic burden of this health problem continues to increase. Research estimates that within the United States, the lifetime cost of domestic violence is approximately $2.1 trillion.[8]

The rates of domestic violence are significantly increased in developing countries in comparison to developed countries.[9] The prevalence of domestic violence in Sub-Saharan Africa (SSA) is above the global average, with a prevalence of approximately 36%.[7] Several African countries rank among some of the highest rates of domestic violence in comparison to other countries.[10] In previous studies, the pooled prevalence of intimate partner violence (IPV) in Sub-Saharan Africa was as high as 44%.[11] Globally, low socioeconomic status is a risk factor for domestic violence, and this remains true in Sub-Saharan African countries.[10] Lower educational attainment also increases one’s risk of experiencing domestic violence during their life.[11]

With this being said, we hypothesize that the political instability and active war that surrounds this area may contribute to this extremely high prevalence of domestic violence. Prior research introduces the idea that active war and violence influences the acceptability and prevalence of violence against women.[12] In addition to this, one study conducted in Liberia suggest that residing in regions with active violence greatly increases the risk of experiencing domestic violence post-conflict.[13]

There is a lack of sufficient and comprehensive research on the prevalence of domestic violence in Africa. Our objective is to determine the prevalence of domestic violence in 30 African countries and to explore the association between domestic violence and wealth, marital status, and educational attainment. In addition to this, we expect that those who are poorer and have less education experience higher rates of domestic violence as well. To our knowledge, this is the first study to estimate the prevalence and incidence of domestic violence in Africa.

## Methods

### Data sources

We used nationally representative Demographic and Health Surveys (DHS) data collected between 2008 and 2018. We extracted data on domestic violence (physical and sexual violence), number of women surveyed, number of households surveyed, education, wealth, and marital status from 30 countries (27 in Sub-Saharan Africa, and 3 from North Africa). We included data from the most recent survey for each country. Surveys without domestic violence were excluded from the analysis. Data collection involved a multistage stratified sampling design, as detailed elsewhere (https://dhsprogram.com/data/data-collection.cfm).

### Ethical considerations

Each country’s ethical review committee and institutional review board of OCF International, USA reviewed and approved the DHS protocol and guideline. Informed consent was collected prior to the survey being given. Data were de-identified and are publically available upon request; therefore, the authors did not need to seek any further clearance.

### Study selection and inclusion criteria

Two authors (AES and PS) independently reviewed and extracted data from each country’s DHS report. Only countries with complete information on physical and sexual violence were included. We derived the following variables: year of the survey, continent, country, number of women surveyed, response rate, proportion of women who experienced sexual violence in the past 12 months, proportion of women who have ever experienced sexual violence, proportion of women who experienced physical violence in the past 12 months, proportion of women who experienced physical violence since age 15, proportion of women abused in each wealth index, proportion of women abused in education level, and proportion of women abused in each marital status. We excluded surveys which were conducted before 2015 or did not report domestic violence data. Non-English reports were translated into English.

### Statistical analysis

The primary outcome was the proportion of women who experienced physical violence within the past 12 months. Secondary outcomes included physical violence since age 15, sexual violence ever, and sexual violence in the past 12 months. We used Poisson regression (meta-regression) to analyze the data. We transformed the proportions into counts by normalizing by offsetting by the number of study participants. All Poisson regression analyses were performed in R studio. Geo-special analysis was conducted using ArcGIS Pro. We performed hot-spot analysis at the sub-regional level using the Getis-ORD Gi* approach with inverse weighting. We calculated G-scores and plotted the results from each outcome and plotted them at the sub-regional level. Information on education, marital status, and wealth was collected and plotted on the country level.

### Patient and public involvement

No patients were involved in the design or dissemination of this analysis.

## Results

We identified 442,670 women from 442,507 households utilizing the DHS data from 2008 to 2018. This set of data represented 30 countries, with 27 of them being located in Sub-Saharan Africa and 3 in Northern Africa. There was a 98% response rate for survey participants, allowing us to utilize a comprehensive pool of data.

### Effects of Education on Domestic Violence

As displaced in **Table 1**, education was significantly associated with physical and sexual violence in a linear fashion, where the most educated had the lowest prevalence and incidence of sexual and physical violence followed by primary, secondary, and higher. Those who had no education were 3.1 times more likely to have experienced physical violence in the last 12 months compared to those of higher education (Risk Ratio: RR= 3.1, 95%CI 3.0-3.16). Likewise, those without any formal education were 2.35 times more likely to have experienced sexual violence within the past 12 months compared to those who had higher education (OR=2.35 95%CI 2.31-2.40).

**Table 1:**
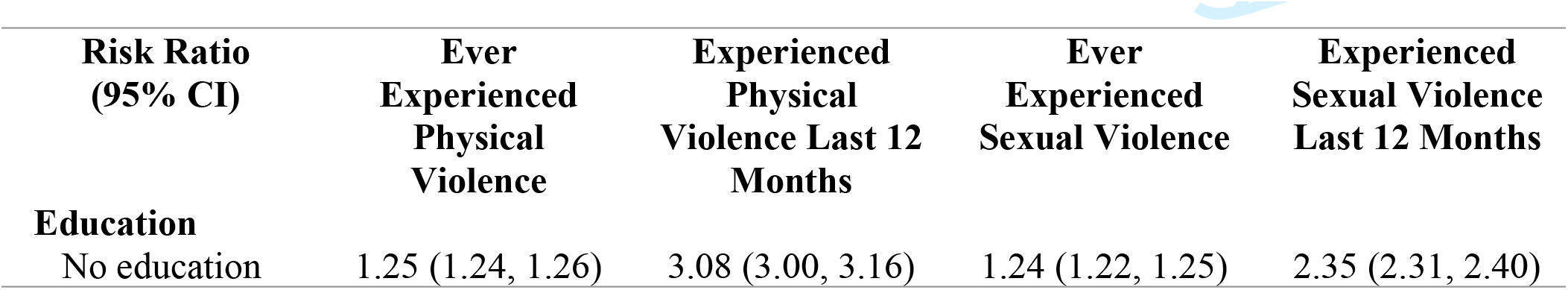
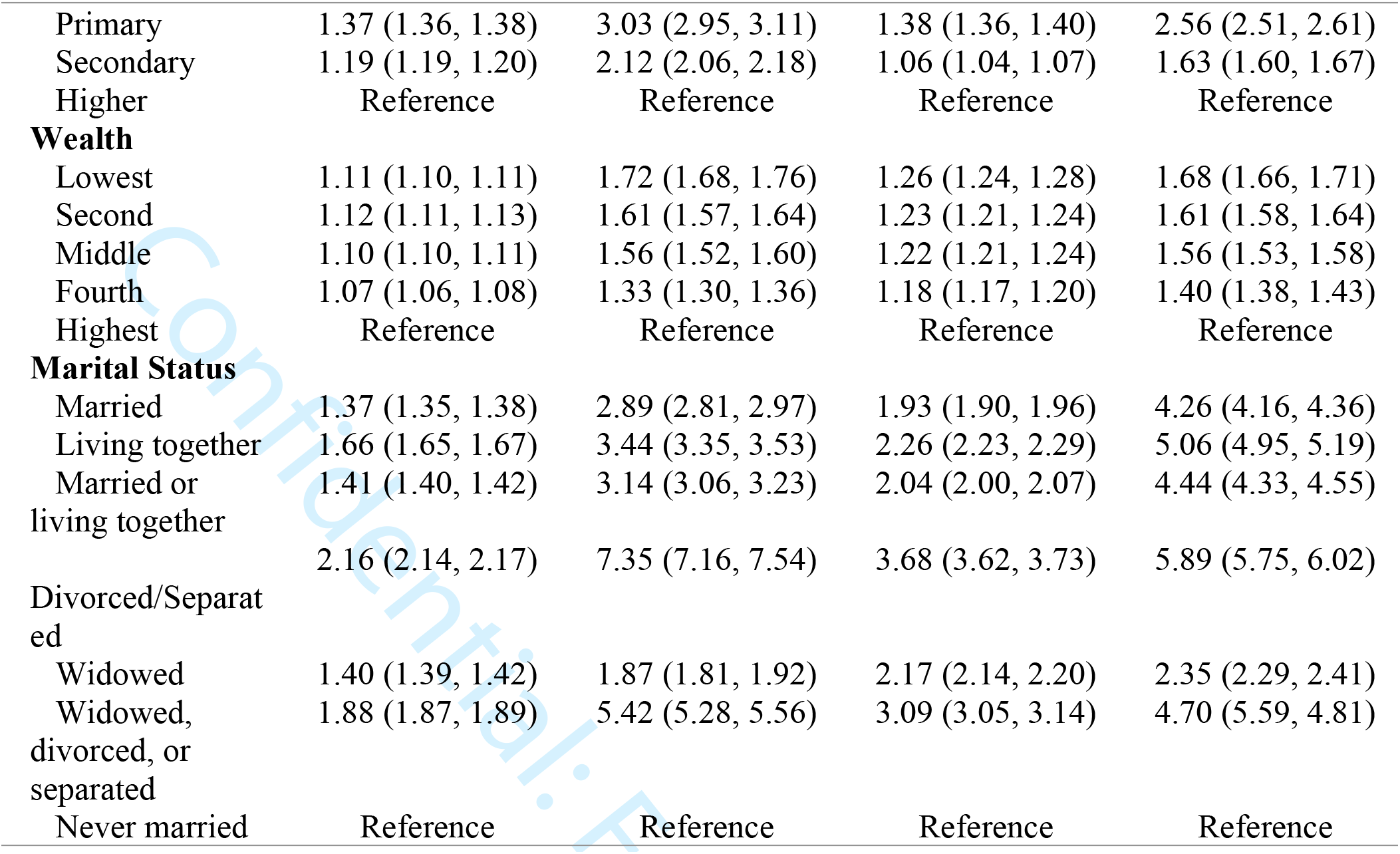

### Effects of Wealth on Domestic Violence

The effects of wealth on the incidence domestic violence appeared linear, where the lowest wealth index was 1.7 times more likely to experience physical violence and 1.68 times more likely to experience sexual violence (RR= 1.72 95%CI 1.68-1.76) and (RR=1.68 95%CI 1.66-1.71) respectfully. As for the prevalence of physical and sexual violence, the lowest wealth index was also at the highest risk for abuse (RR=1.11 95%CI 1.1-1.11) and (RR=1.26 95%CI = 1.24-1.28). Effects of Marital Status on Domestic Violence

### Effects of Marital Status on Domestic Violence

Those who were divorced or separated were 7.5 times more likely to have experienced physical violence in the past 12 months (RR= 7.35, 95%CI 7.16-7.54) and 5.9 times more likely to have experienced sexual violence in the past 12 months (RR-5.89, 95%CI=5.75-6.02) compared to those who were never married. Living together also increased the incidence rate of physical and sexual violence by 3.4 and 5.1 times, compared to those never married, respectfully

### Geospatial Statistics

Sub-regions near Congo had the highest prevalence and incidence of physical and sexual violence (**Figure 1**). Many regions in this area were identified as hotspots (**Figure 2**). Areas in northern Africa had lower incidence and prevalence rates of sexual and physical violence. Many regions in northern Africa were identified as cold spots.

**Figure 1:**
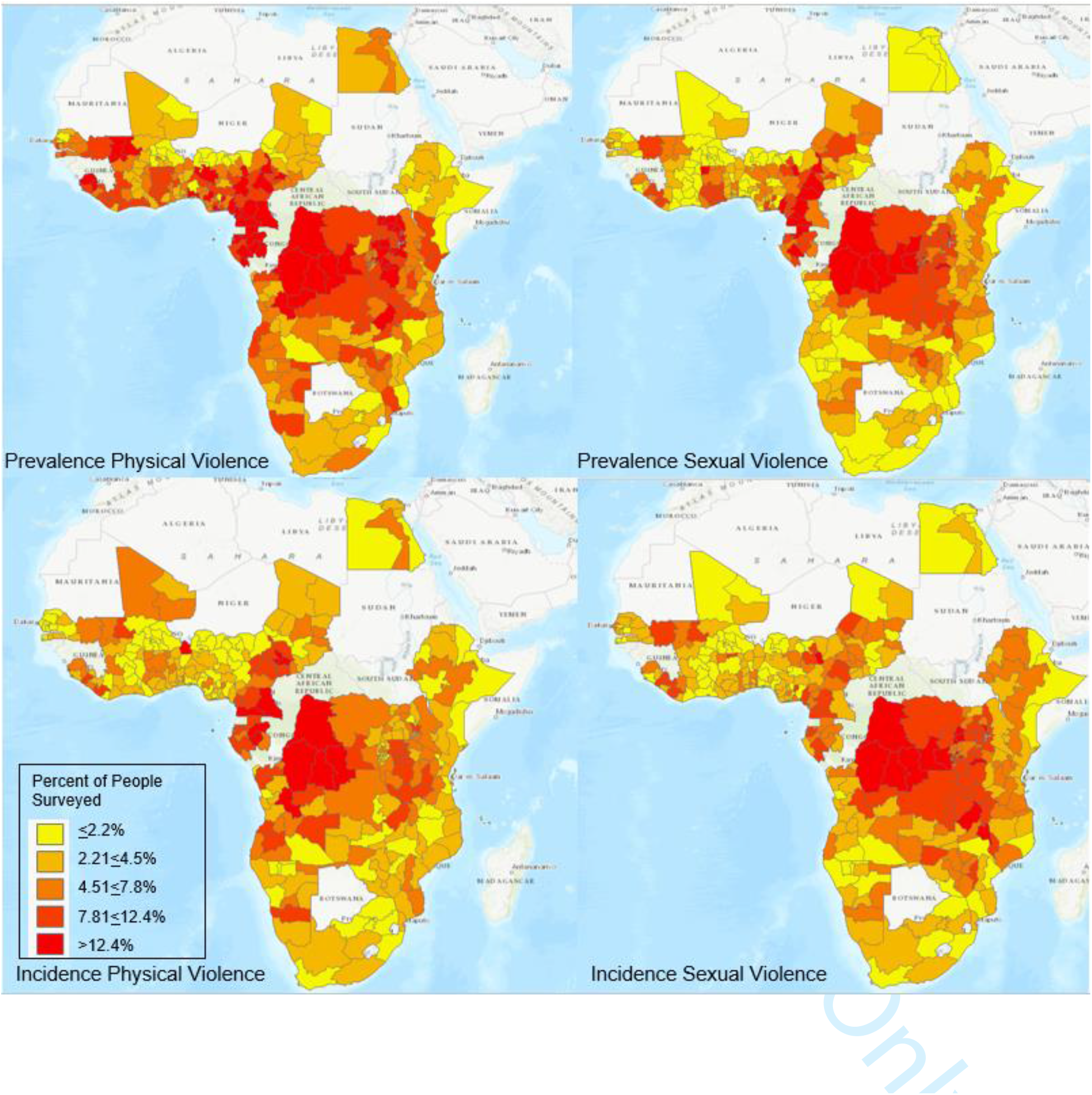
Prevalence and incidence of physical and sexual violence in 30 countries in Africa

**Figure 2:**
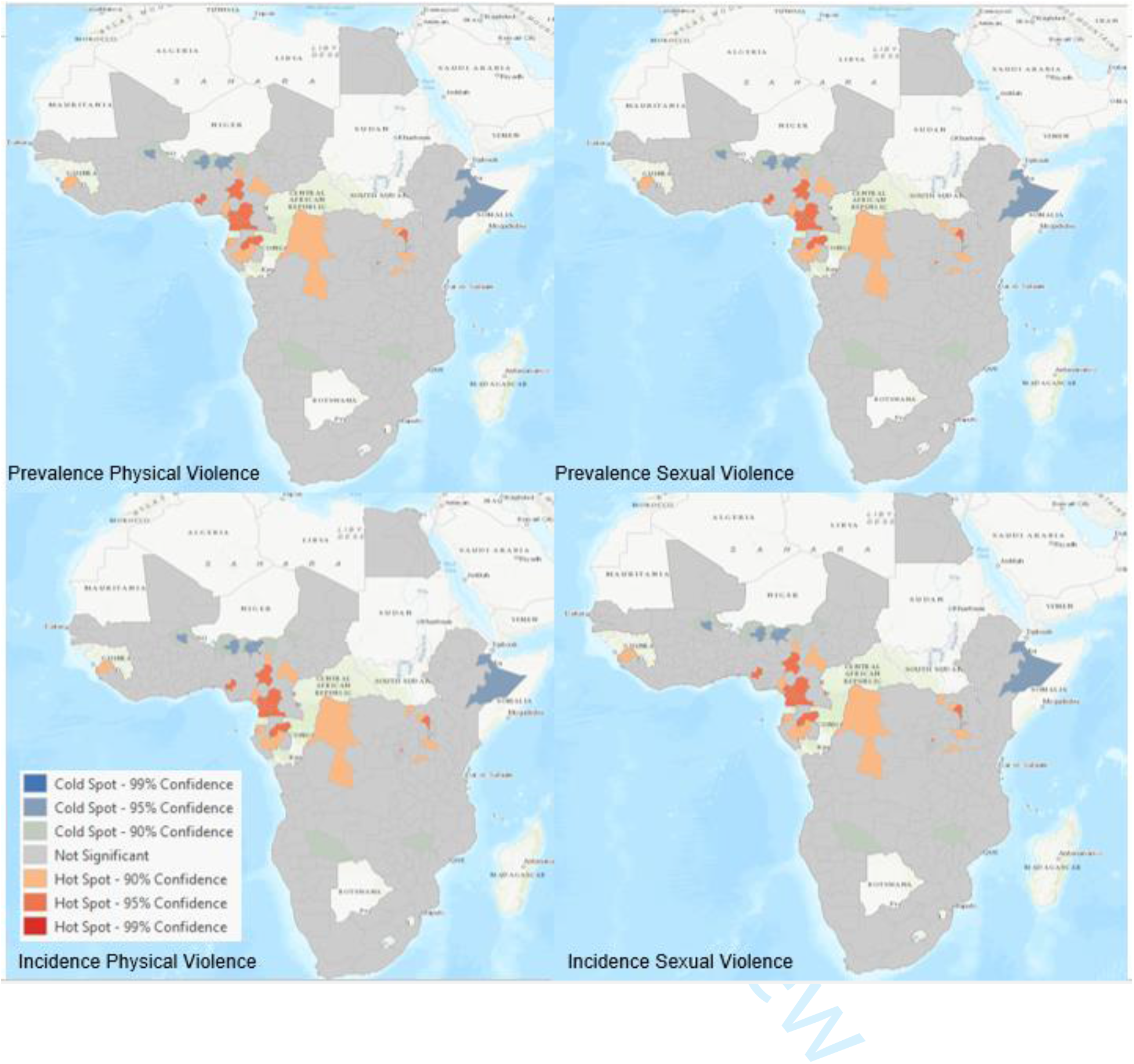
Hotspots for physical and sexual violence in 30 countries in Africa

## Discussion

We utilized data from the Demographic and Health Surveys (DHS) to estimate the prevalence of domestic violence and how wealth, marital status, and educational attainment affects the rates of physical and sexual violence. To our knowledge, this is the first comprehensive ecological study to look at the prevalence of domestic violence in 30 African regions and to explore the association between domestic violence and socioeconomic status, educational attainment, and marital status. Our results suggest that sub-regions located in close proximity to the Democratic Republic of Congo (DRC) had the highest rates of both physical and sexual violence. On the contrary, as seen on the geospatial map, northern Africa had the lowest rates of domestic violence. The cold spots seen in these maps represent areas with a decreased prevalence of physical and sexual violence. These findings are in line with previous research, which suggests that Sub-Saharan African countries have an above average prevalence of domestic violence in comparison to other African regions.[7]

When analyzing the association between socioeconomic status, it was found that those in the lowest wealth category are most likely to experience both physical and sexual violence. There is an inverse relationship between wealth and risk of domestic violence, meaning that as wealth decreases, the risk of both physical and sexual violence increases simultaneously. Those in the lowest wealth category were 1.7 times (RR 1.72; 95% CI 1.68, 1.76) more likely to experience physical violence in the past 12 months and 1.68 times (RR 1.68; 95% CI 1.66, 1.71) more likely to experience sexual violence in the past 12 months. Overall, the lowest wealth category was 1.11 times (RR 1.11; 95% CI 1.1, 1.11) more likely to ever experience physical violence and 1.26 times (RR 1.26; 95% CI 1.24, 1.28) more likely to ever experience sexual violence in their lifetime. This was followed by those in the second wealth category, who were 1.61 times more likely to experience both physical (RR 1.61; 95% CI 1.57, 1.64) and sexual violence (RR 1.61; 95% CI 1.66, 1.71) in the past 12 months. Those categorized in the middle wealth category were 1.56 times more likely to experience both physical (RR 1.56; 95% CI 1.52, 1.6) and sexual violence (RR 1.56; 1.53, 1.58) in the past 12 months. Individuals in this group were 1.1 times (RR 1.1; 95% CI 1.10, 1.11) more likely to ever experience physical violence and 1.22 (RR 1.22; 95% CI 1.21, 1.24) times more likely to ever experience sexual violence. On the contrary, those in the highest wealth categories, were least likely to have experienced any physical or sexual violence at any time in comparison to lower wealth categories. The relationship between socioeconomic status and domestic violence risk is one that has been founded in evidence. Therefore, our findings are in line with previous research that support the notion that individuals in a lower socioeconomic status bracket are more likely to experience any form of physical or sexual violence.[14]

The association between educational attainment and risk of domestic violence was also inverse. Those with lower levels of education were more likely to have experienced physical and sexual violence at any time. Those with no education were 3.1 times (RR 3.08; 95% CI 3.00, 3.16) more likely to have experienced physical violence in the past 12 months and 2.35 times (RR 2.35; 95% CI 2.31, 2.4) more likely to experience sexual violence in the past 12 months. Those with a primary education were 3.03 times (RR 3.03; 95% CI 2.95, 3.11) more likely to have experienced physical violence in the past 12 months and 2.56 times (RR 2.56; 95% CI 2.51, 2.61) more likely to have experienced sexual violence in the past 12 months. This group is 1.37 times (RR 1.37; 95% CI 1.36, 1.38) more likely to have ever experienced physical violence and 1.38 times (RR 1.38; 95% CI 1.36, 1.40) more likely to have ever experienced sexual violence. Those with a secondary education experience the lowest risk of physical and sexual violence. This group is 2.12 times (RR 2.12; 95% CI 2.06, 2.18) more likely to have experienced physical violence in the past 12 months and 1.63 times (RR 1.63; 95% CI 1.60, 1.67) more likely to have experienced sexual violence in the past 12 months. In addition to this, those with a secondary education are 1.19 times (RR 1.19; 95% CI 1.19, 1.20) more likely to have ever experienced physical violence and 1.06 times (RR 1.06; 95% CI 1.04, 1.07) more likely to have ever experienced sexual violence in their lifetime.

Those who are divorced/separated have the highest risk of both physical and sexual violence at any point. These individuals are 7.35 times (RR 7.35; 95% CI 7.16, 7.54) more likely to have experienced physical violence in the past 12 months and 5.89 times (RR 5.89; 95% CI 5.75, 6.02) more likely to have experienced sexual violence in the past 12 months. They also have the highest risk of both physical and sexual violence at any point in time. Those who are divorced or separated are 2.16 times (RR 2.16; 95% CI 2.14, 2.17) more likely to experience physical abuse and 3.68 times (RR 3.68; 95% CI 3.62, 3.73) more likely to experience sexual violence at any time. This is followed by individuals who cohabitate, or live together. Those who are living together are 3.44 times (RR 3.44; 95% CI 3.35, 3.53) more likely to have experienced physical violence in the past 12 months and 5.06 times (RR 5.06; 95% CI 4.95, 5.19) more likely to have experienced sexual violence in the past 12 months. Those who have never been married or are single experience the lowest risk of domestic violence.

## Conclusions

To summarize, domestic violence was most common in Democratic Republic of Congo and surrounding countries. Wealth and educational attainment are both inversely related to risk of domestic violence. Those who are divorced experience an increased risk of domestic violence, followed by individuals living together, then married individuals. Single or non-married individuals experience the lowest risk of domestic violence.

Public health interventions targeted at domestic violence and the adverse consequences that may arise as a result of this experience are continuously growing and expanding. Prior research suggests that therapeutic interventions may be effective in addressing the negative effects of domestic violence.[15] With this being said, implementing interventions in African regions focused on the adverse consequences of domestic violence may be useful. While the evidence of if specific interventions are effective in reducing the prevalence of domestic violence are inconclusive, further research is necessary to explore potential ways to reduce these growing and concerning rates of violence.

## Data Availability

All data is publicly available in the Demographic and health survey (DHS) published reports.

https://dhsprogram.com/

## Contributors

AS, PS, JSO, and JH conceived the study. AS, EH, and YY conducted the literature search. AS, DL, VC, and PS completed data analysis. AS, DL, VC, JY, PS, JSO, JO and JH interpreted the data. AS, ES, YY wrote the manuscript. All Authors agreed to the manuscript in its final form

